# A machine learning-based model for survival prediction in patients with severe COVID-19 infection

**DOI:** 10.1101/2020.02.27.20028027

**Authors:** Li Yan, Hai-Tao Zhang, Jorge Goncalves, Yang Xiao, Maolin Wang, Yuqi Guo, Chuan Sun, Xiuchuan Tang, Liang Jin, Mingyang Zhang, Xiang Huang, Ying Xiao, Haosen Cao, Yanyan Chen, Tongxin Ren, Fang Wang, Yaru Xiao, Sufang Huang, Xi Tan, Niannian Huang, Bo Jiao, Yong Zhang, Ailin Luo, Laurent Mombaerts, Junyang Jin, Zhiguo Cao, Shusheng Li, Hui Xu, Ye Yuan

## Abstract

The sudden increase of COVID-19 cases is putting a high pressure on healthcare services worldwide. At the current stage, fast, accurate and early clinical assessment of the disease severity is vital. To support decision making and logistical planning in healthcare systems, this study leverages a database of blood samples from 404 infected patients in the region of Wuhan, China to identify crucial predictive biomarkers of disease severity. For this purpose, machine learning tools selected three biomarkers that predict the survival of individual patients with more than 90% accuracy: lactic dehydrogenase (LDH), lymphocyte and high-sensitivity C-reactive protein (hs-CRP). In particular, relatively high levels of LDH alone seem to play a crucial role in distinguishing the vast majority of cases that require immediate medical attention. This finding is consistent with current medical knowledge that high LDH levels are associated with tissue breakdown occurring in various diseases, including pulmonary disorders such as pneumonia. Overall, this paper suggests a simple and operable formula to quickly predict patients at the highest risk, allowing them to be prioritised and potentially reducing the mortality rate.

**Funding:** None.

## Introduction

The outbreaks of COVID-19 epidemic have been causing worldwide health concerns since December 2019. The virus causes fever, cough, fatigue and mild to severe respiratory complications, eventually leading to patient death. On March 6, the total amount of cumulated infection cases over the world was 97,000 and 3,400 deaths [WHO]. On March 11, the virus outbreak was declared a pandemic by the World Health Organization, as the virus spread to 114 countries, totalling over 118,000 recorded cases and 4,300 deaths [WHO]. So far, it has been reported that 26.1-32.0% of COVID-19 infected patients are prone to develop critically ill cases^1,2^. Furthermore, recent reports expose an astonishing case fatality rate of 61.5% for critical cases, increasing sharply with age and for patients with underlying comorbidities^3^. Both the reach and severity of cases is putting great pressure on the medical services, and readily lead to a shortage of intensive care resources.

Unfortunately, so far, there is currently no available prognostic biomarker to distinguish patients that require immediate medical attention, and their associated mortality rate. The capacity to distinguish cases that are at imminent risk of mortality, therefore, has become an urgent yet challenging necessity. Under those circumstances, we retrospectively analysed blood samples of 404 patients from the region of Wuhan, China to identify robust and meaningful markers of mortality risk. For this purpose, a mathematical modelling approach based on state-of-the-art interpretable machine learning algorithms was devised to identify the most discriminative biomarkers of patient survival. The problem is formulated as a classification task, where the inputs are basic information, symptoms, blood samples, laboratory test, including liver function, kidney function, coagulation function, electrolyte, inflammatory factors taken from originally general, severe and critical patients (Table 1) and their associated outcomes corresponding to either survival or death at the end of the examination period. Through optimization, the classifier aims to reveal the most crucial biomarkers distinguishing patients at imminent risk, thereby relieving clinical burden and potentially reducing the mortality rate.

**Table 1:**
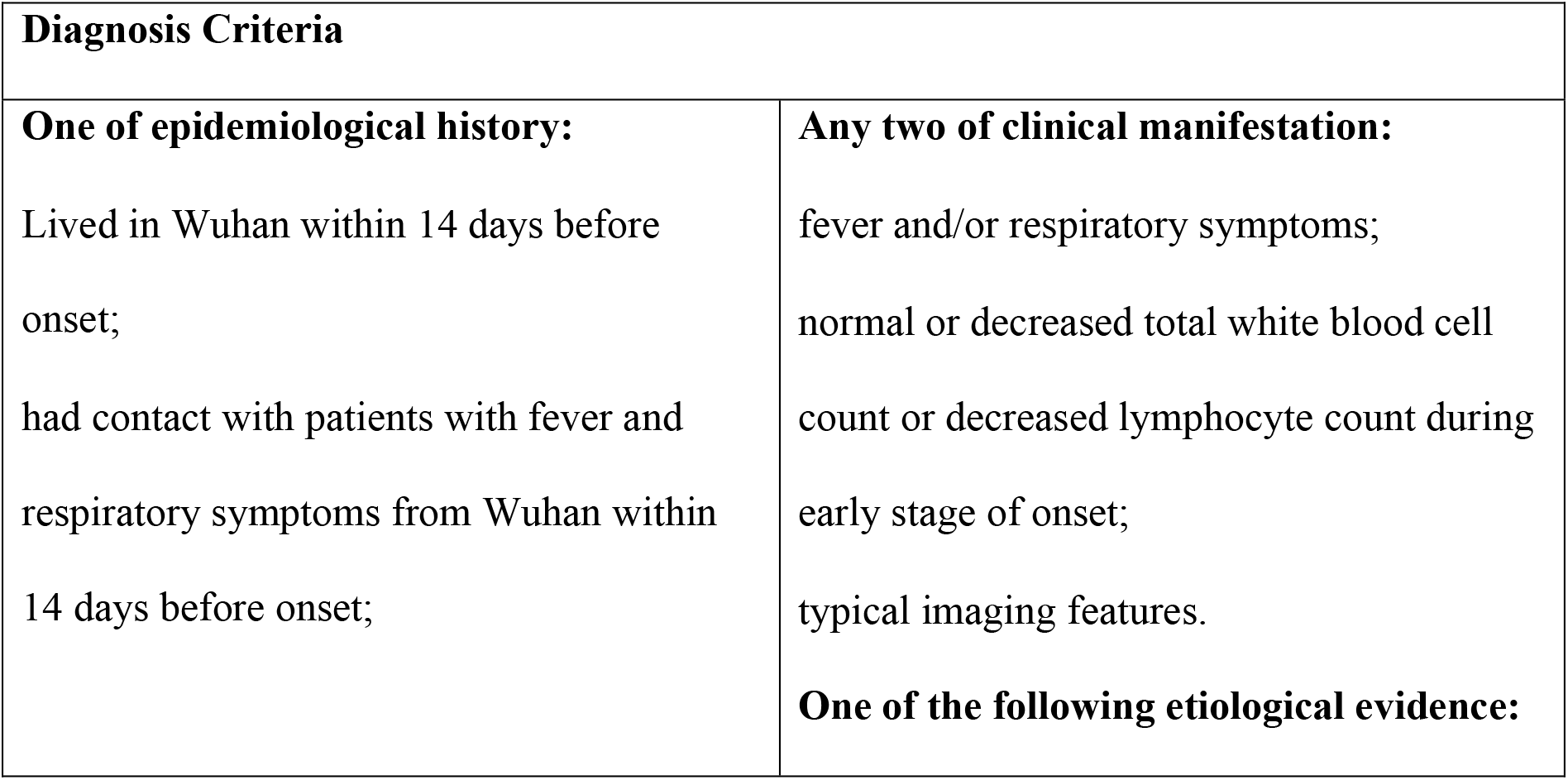
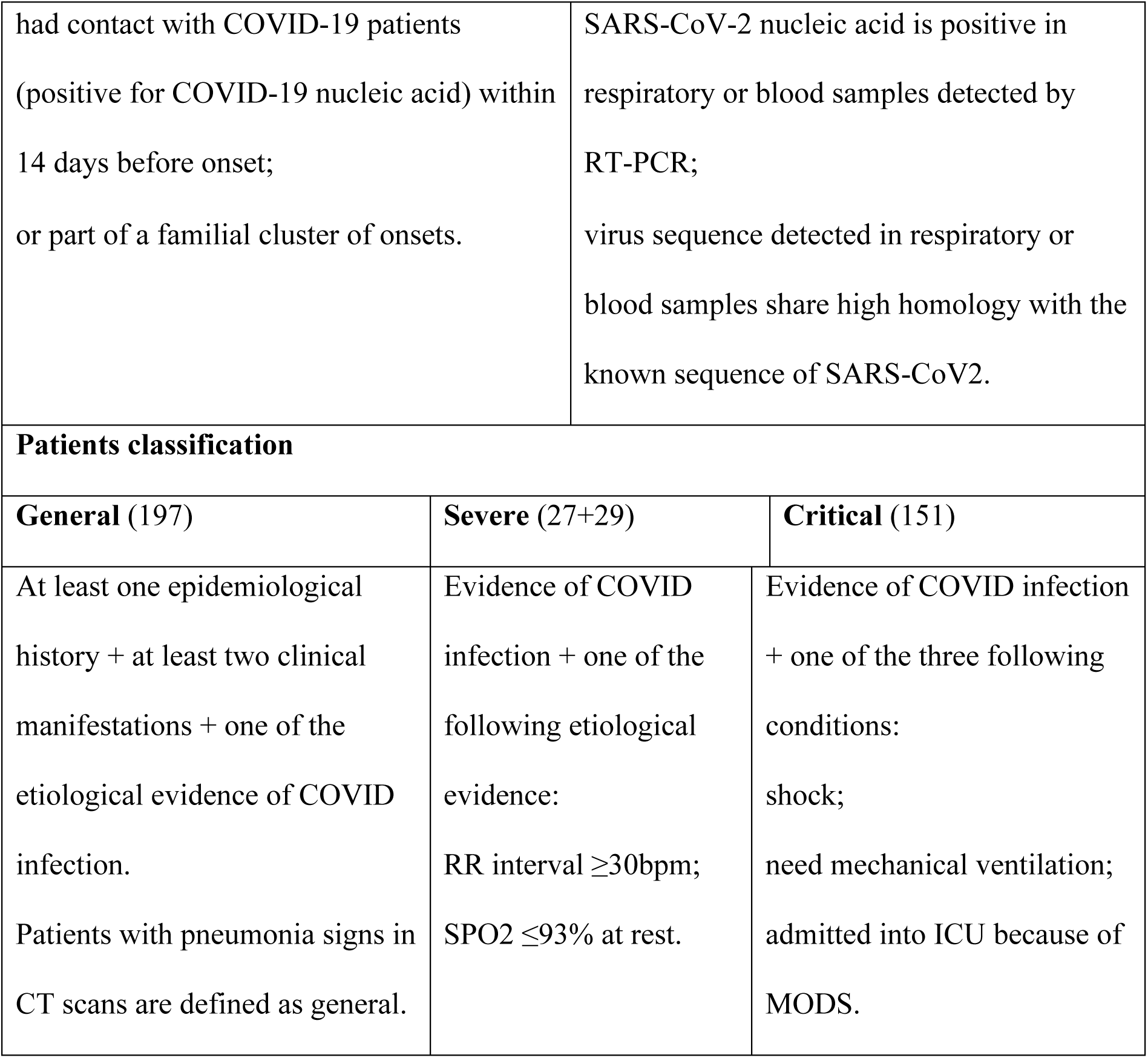
Criteria for assessment of disease severity upon Hospital admission.

| **Diagnosis Criteria** |  |
| --- | --- |
| **One of epidemiological history:**<br>Lived in Wuhan within 14 days before onset;<br>had contact with patients with fever and respiratory symptoms from Wuhan within 14 days before onset; | **Any two of clinical manifestation:**<br>fever and/or respiratory symptoms;<br>normal or decreased total white blood cell count or decreased lymphocyte count during early stage of onset;<br>typical imaging features.<br>**One of the following etiological evidence:** |

|  |  |  |
| --- | --- | --- |
| had contact with COVID-19 patients (positive for COVID-19 nucleic acid) within 14 days before onset;<br>or part of a familial cluster of onsets. |  | SARS-CoV-2 nucleic acid is positive in respiratory or blood samples detected by RT-PCR;<br>virus sequence detected in respiratory or blood samples share high homology with the known sequence of SARS-CoV2. |
| **Patients classification** |  |  |
| **General (197)** | **Severe (27+29)** | **Critical (151)** |
| At least one epidemiological history + at least two clinical manifestations + one of the etiological evidence of COVID infection.<br>Patients with pneumonia signs in CT scans are defined as general. | Evidence of COVID infection + one of the following etiological evidence:<br>RR interval $\geq 30$bpm;<br>SPO2 $\leq 93\%$ at rest. | Evidence of COVID infection + one of the three following conditions:<br>shock;<br>need mechanical ventilation;<br>admitted into ICU because of MODS. |

Medical records were collected using standard case report forms that included epidemiological, demographic, clinical, laboratory, drugs, nursing, and mortality outcome (Table 2). The clinical outcomes were followed up to 20 February 2020. The study was approved by the Tongji Hospital Ethics Committee.

**Table 2:**
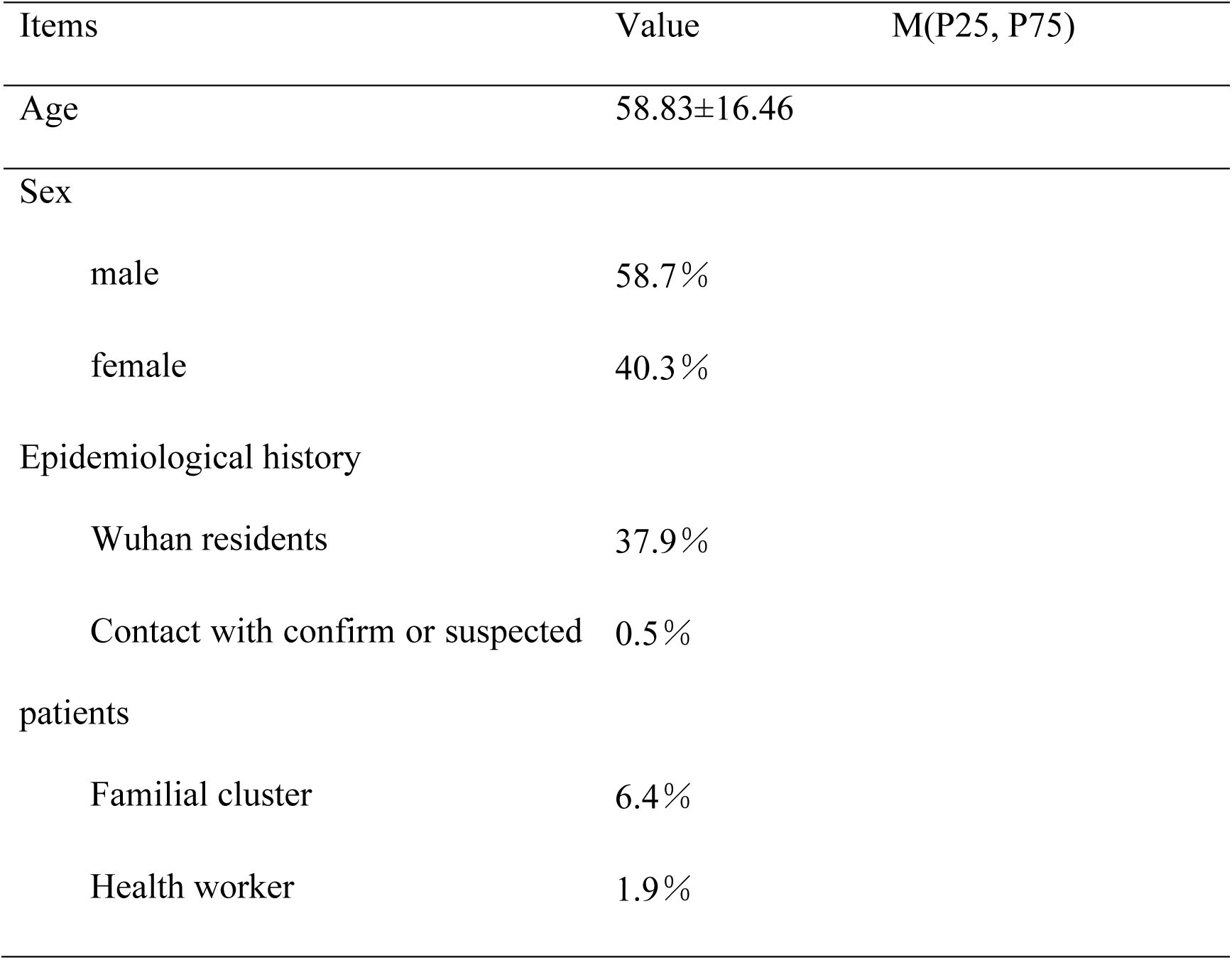
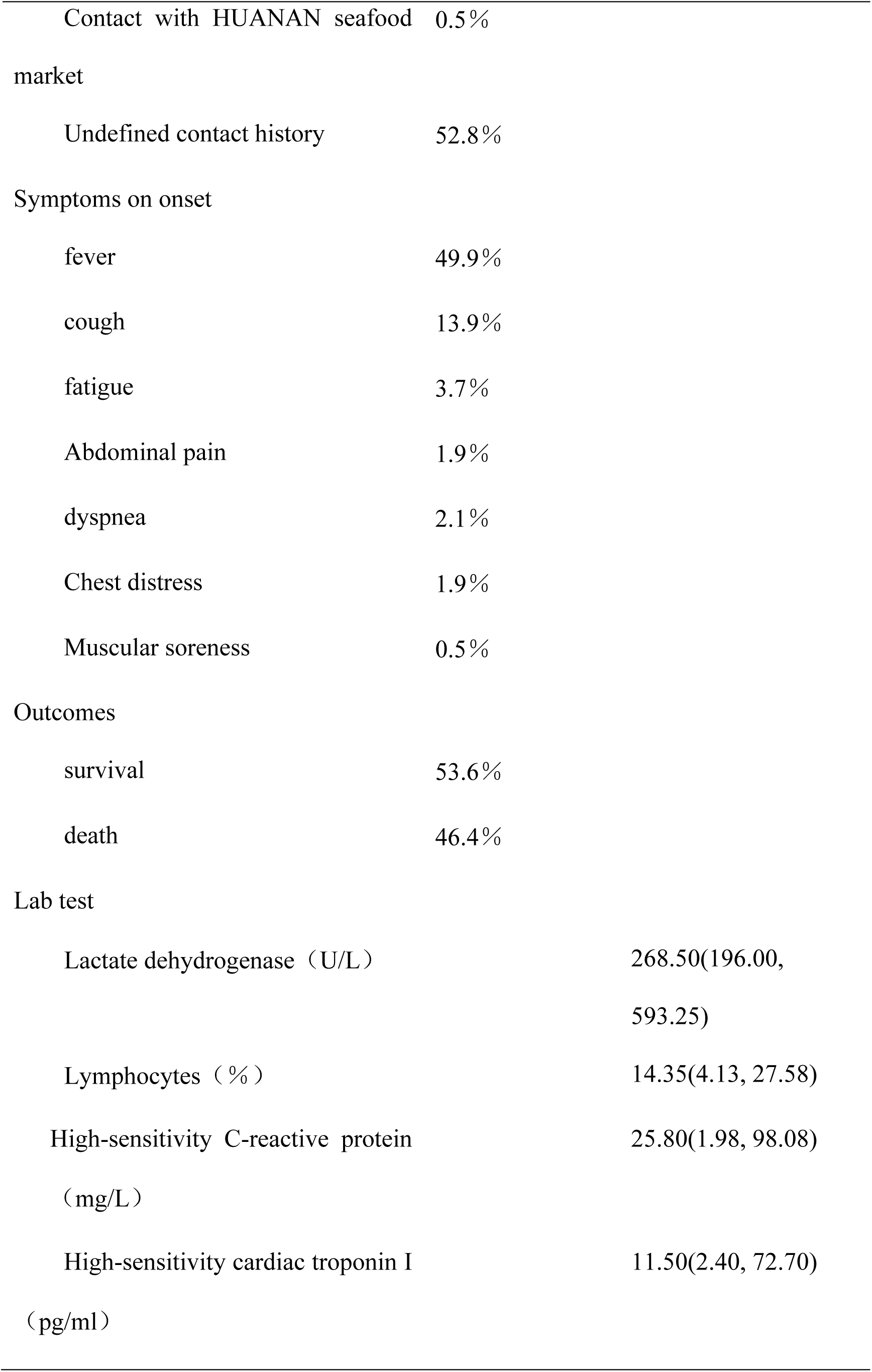
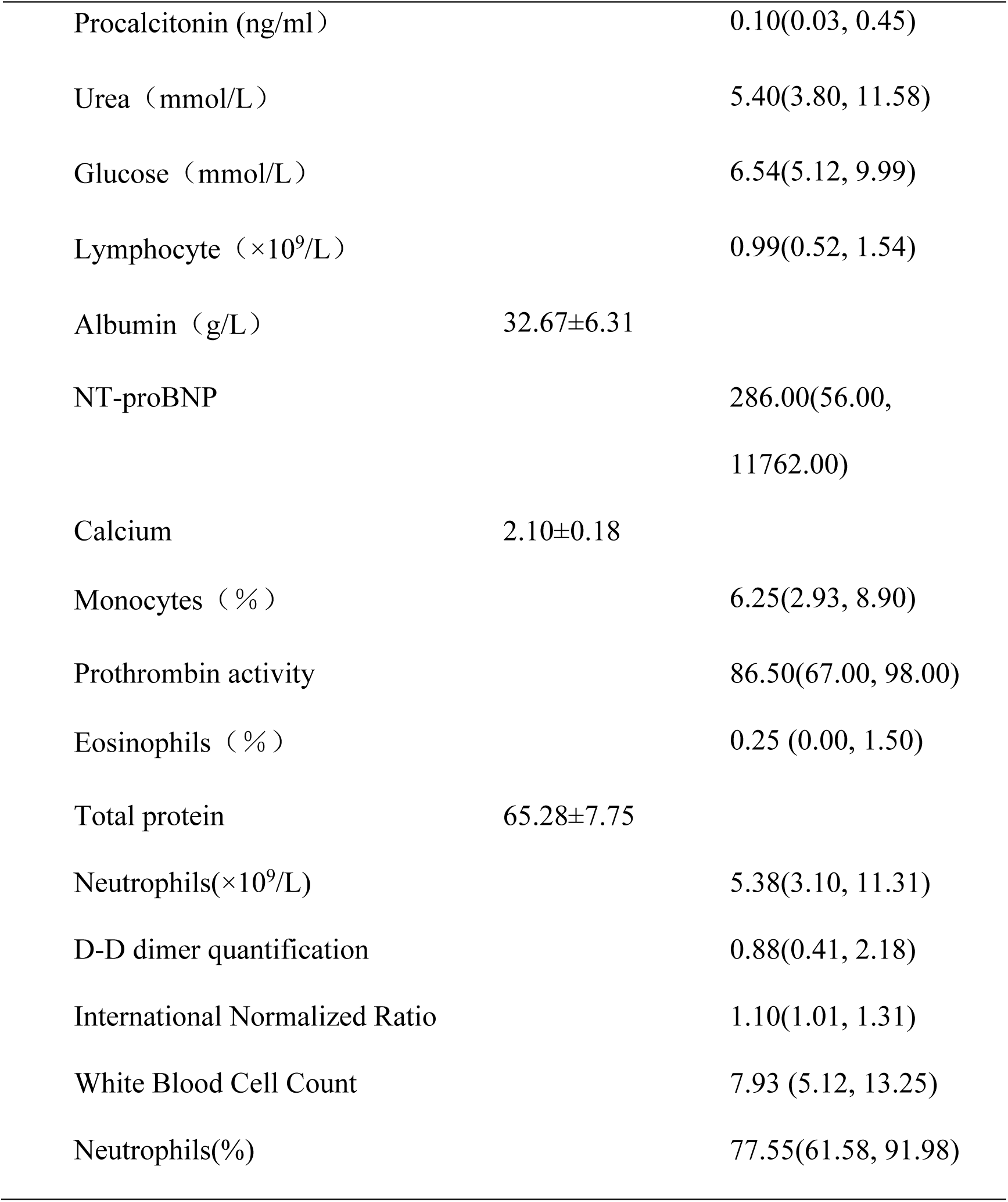
the continuity variables of normal distributions were described by mean ± standard deviation, and the continuity variables of non-normal distributions were described by median and quartile. Data were first tested for normality. The Kolmogorov-Smirnov test (K-S test for short) tested whether a single sample is from a particular distribution; then, this single sample K-S test checked the normality of data. A test level of α=0.05, and P <0.05 indicate that a sample does not fit a normal distribution. Since age, total protein, albumin, and calcium all fitted normal distributions, only their means ± standard deviations were used to describe their concentration trend. For all other continuous variables (fitting non-normal distributions), median described their concentration trend.

### Statistical Analysis of Electronical Records

We considered the medical information of all patients collected between 10 January and 20 February 2020. Data originating from pregnant and breast-feeding women, patients younger than 18 years old, and recordings without at least 80% of complete data materials, were excluded from subsequent analysis. Out of the 404 remaining patients, 213 recovered from the virus, while the remaining 191 died. This high mortality rate is related to the fact that Tongji Hospital admitted most severe cases in Wuhan. Upon admission, patient’s severity was empirically assessed by medical doctors according to the rules in Table 1^4^. Figure 1A summarises the outcome of patients in the three different classifications.

**Figure 1:**
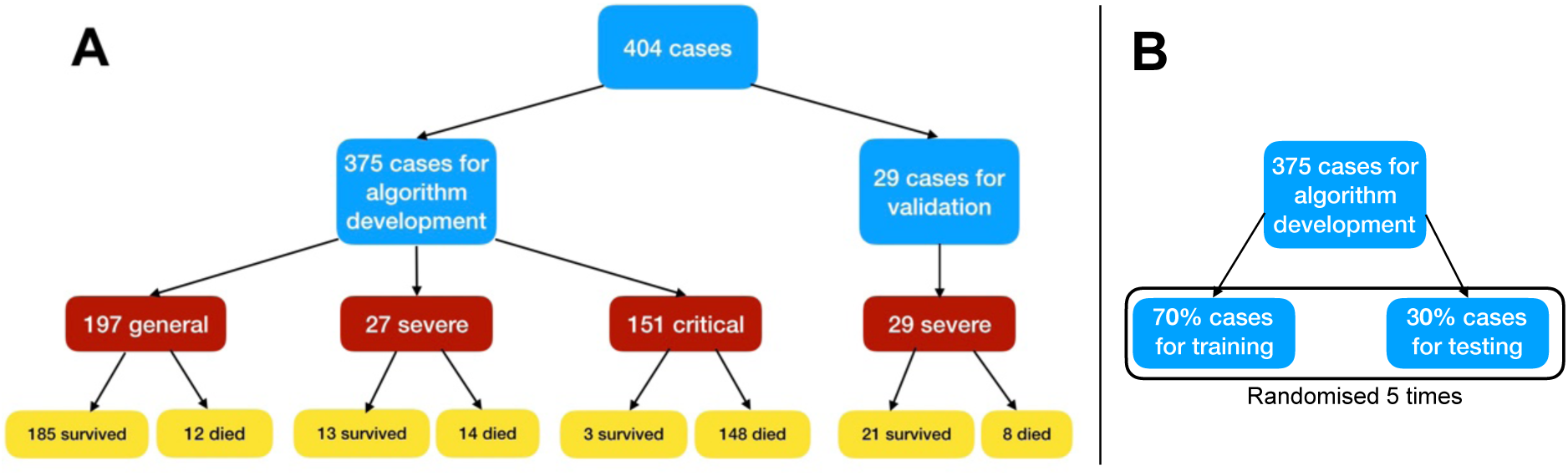
**A**: A flowchart of patient enrollment. **B**: Dividing of data for training and testing.

Data of 404 patients were recovered from Excel 2016, and double checked through SPSS 26.0 analysis data. Then, patients’ data were separated into training, test and additional validation sets. Training and test sets altogether considered 375 patients, while the validation set consisted of 29 patients (Figure 1). The validation set was chosen with only severe patients since these are the most unpredictable in terms of clinical outcome. Table 2 displays the statistics of blood samples for those patients. Fever was the most common initial symptom (49.9%), followed by cough (13.9%), fatigue (3.7%), and dyspnoea (2.1%). The age distribution of the 375 patients was 58.83 ± 16.46 years old, with 58.7% of males. The epidemiological history included Wuhan residents (37.9%), familial cluster (6.4%), and health workers (1.9%).

#### Model Training

While most patients were taken multiple blood samples throughout their stay at the hospital, the model training and testing uses only the last available recordings of patients as inputs to the model to assess crucial biomarkers of disease severity, distinguish patients that require immediate medical assistance, and accurately matching corresponding features to each label. Missing data were “–1” padded. The model output corresponds to patient survival. Patients that survived were assigned to class 0, and those that died to class 1.

The 375 cases used to develop the model were first separated into a training and a test set following a 7:3 ratio, further cross-validated 5 times (Figure 1B). The performance models were evaluated by assessing the classification accuracy (ratio of true predictions over all predictions), the precision, sensitivity/recall and F1 scores (defined below).

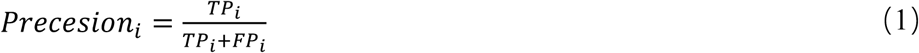

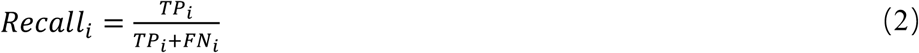

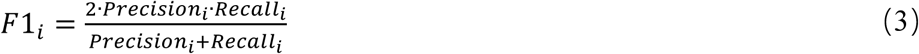

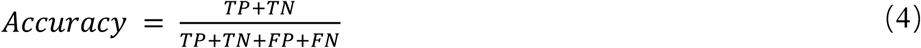

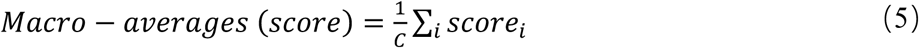

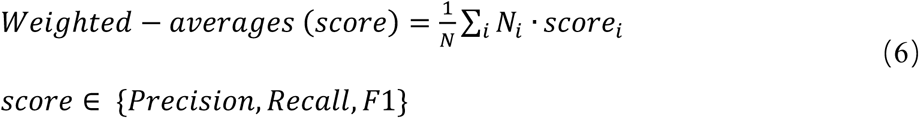

where *i* ∈ *C* represents the class, N is the number of all samples, *N*_*i*_ is the number of samples, TN_*i*_ in class *i*, TP_*i*_, FP_*i*_, and FN_*i*_ stand for true positive, true negative, false positive and false negative rates for class *i*, respectively. In total, 84 features were considered.

This study uses a supervised XGBoost classifier^5^ as the predictor model. XGBoost is a high-performance machine learning algorithm that benefits from great interpretability potential due to its recursive tree-based decision system. In contrast, internal model mechanisms of_black-box modelling strategies are typically_difficult to interpret. The importance of each individual feature in XGBoost is determined by its accumulated use in each decision step in trees. This computes a metric characterizing the relative importance of each feature, which is particularly valuable to estimate features that are the most discriminative of model outcomes. Especially, when they are related to meaningful clinical parameters.

### Model Optimization

XGBoost was originally trained with default parameter settings: max depth equal to 4, learning rate equal to 0.2, number of tree estimators set to 150, value of the regularization parameter α set to 1 and ‘subsample’ and ‘colsample_bytree’ both set to 0.9 to prevent overfitting for cases with many features and small sample size^5^.

#### Feature Selection

To evaluate markers of imminent mortality risk, we assessed the contribution of each patient parameters to decisions of the algorithm through a feature selection process. Features were ranked by XGBoost according to their importance (Supplementary Figure 1). Essentially, the algorithm, detailed in Supplementary Algorithm 1, selected three features. Supplementary Figure 2 shows no performance improvement in F1-scores when the number of top features increased to 4. Hence, the number of key features was set to the following 3: LDH, lymphocytes and hs-CRP.

Table 3 displays the performances of the model for both the train/test splits and the additional validation set. The results show that the model is able to accurately predict the outcome of patients, regardless of their original diagnosis upon Hospital admission. Moreover, the performance of the additional validation set is similar to that of the training and test sets. This suggests that the model captures the key biomarkers for patient survival. The table further emphasizes the importance of LDH as a crucial biomarker for patient survival rate.

**Table 3:** performances of the XGBoost classification in discriminating between mortality outcomes.

| Features | Cross-validation F1-score<br>(mean +- std) | Additional<br>validation set F1-score (mean +- std) |
| --- | --- | --- |
| LDH | 93.85% +- 0.87% | 91.69% +- 2.88% |
| LDH, Lymphocyte | 96.43% +- 0.48% | 93.98% +- 2.44% |
| LDH, Lymphocyte, hs-CRP | 97.59% +- 0.40% | 94.67% +- 2.34% |

### Explainable model

Following previous findings on the importance of LDH, lymphocytes and hs-CRP, the next step is to construct a simplified and clinically operable decision model. XGBoost algorithms are based on recursive decision tree building from past residuals and can identify those trees that contribute the most to the decision of the predictive model. Decision trees are simple classifiers consisting of sequences of binary decisions organized hierarchically. Single decision trees are intuitively appealing since they are based on a recursive dichotomic partitioning of the data following an optimal separating decision rule at each node. Hence, if the accuracy of a tree remains high, reducing the complexity of the model to such structure has the potential to reveal a clinically portable decision algorithm. Below, we further refer to the latter as “explainable model”, or “single-tree XGBoost”.

There was a total of 24 patients with incomplete measurements for at least one of the three principal biomarkers, leaving 351 patients to identify a single-tree XGBoost model. To identify such a model, XGBoost is re-trained with the same parameters as described above, except: number of tree estimators set to 1, values of the regularization parameters α and β both set to 0, and the subsample and max features both are set to 1 as over-fitting issues have been avoided based on previous modelling^5^. The algorithm computed accuracy, precision, sensitivity, recall and F1 scores on the test samples and additional validation set. The best performing tree chosen based on test datasets, together with its accuracy, is shown in Figure 2. Supplementary Tables 1 and 2 show performance of the Multi-tree XGBoost algorithm on the training and testing datasets, with accuracies of 96% and 97%, respectively.

**Figure 2.**
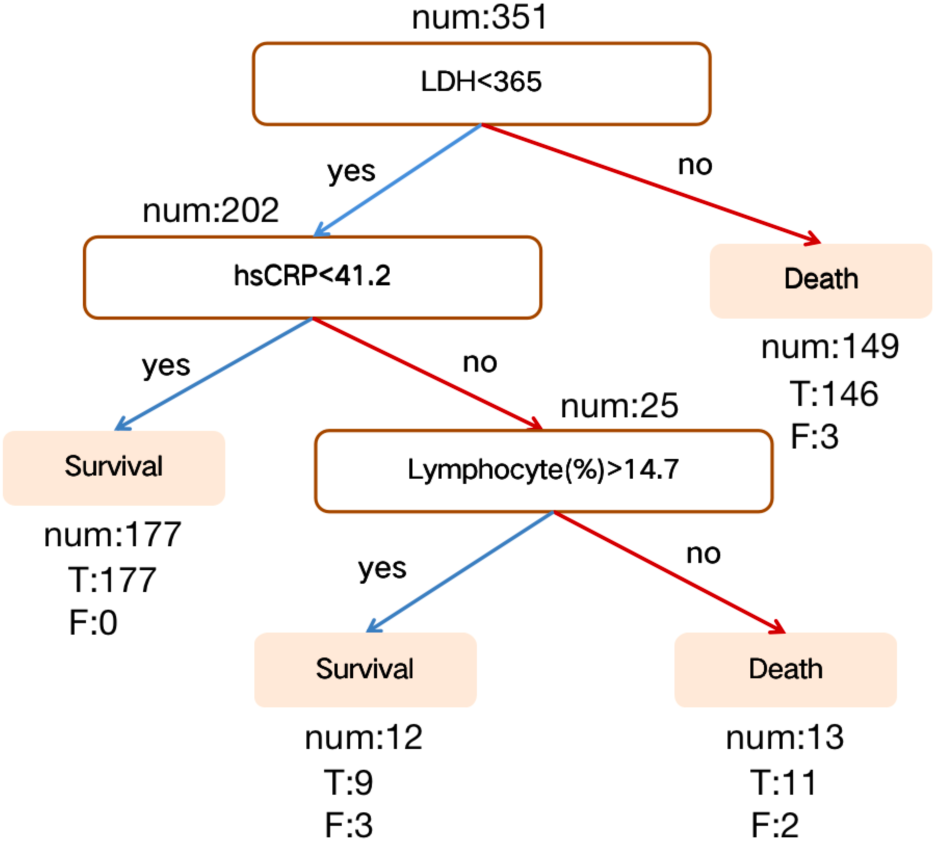
Discovered a decision rule using three key features and their threshold in absolute value. Num represents the number of patients in a class, T represents the number of corrected classified while F represents the number of misclassified patients.

The performance of the model on the external validation data, which has not been part of training or testing the model, can be found in Table 4. Its associated confusion matrix is shown on the left of Figure 3, showing 100% death prediction accuracy and 90% survival prediction accuracy. The confusion matrix on the right of Figure 3 corresponds to the latest measurements of 375 patients. Overall, scores for survival and death prediction, accuracy, macro and weighted averages are consistently over 0.90. Remarkably, both Multi-tree XGBoost and Single-tree XGBoost return the similar Predictions (Supplementary Figure 3).

**Table 4:** Performance of the proposed algorithm on testing dataset.

|  | Precision | Recall | F1-score | Support |
| --- | --- | --- | --- | --- |
| Survival | 1.00 | 0.83 | 0.91 | 12 |
| Death | 0.89 | 1.00 | 0.94 | 17 |
| accuracy |  |  | 0.93 | 29 |
| macro avg | 0.95 | 0.92 | 0.93 | 29 |
| weighted avg | 0.94 | 0.93 | 0.93 | 29 |

**Figure 3:**
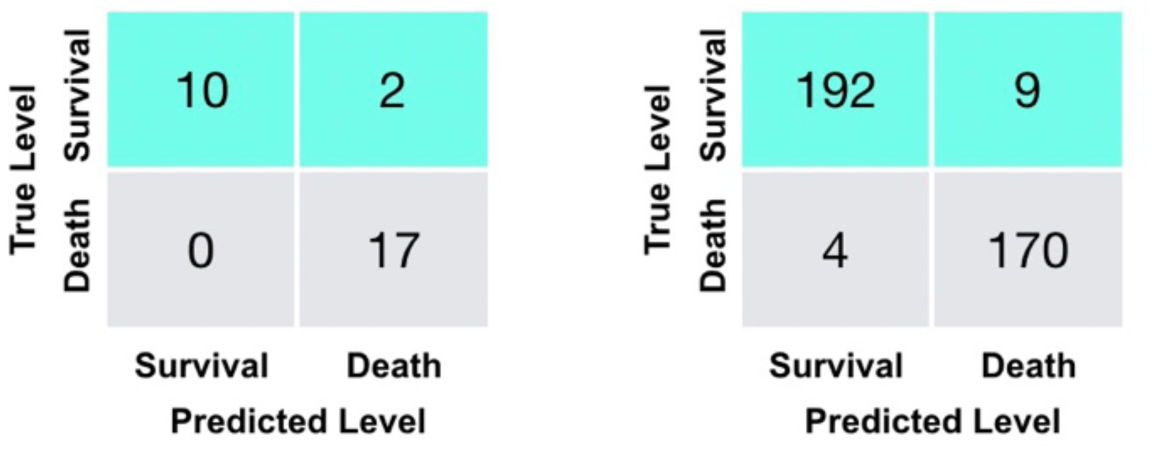
Confusion matrices for the additional validation set (29 severe patients) (left) and training and testing sets (375 patients) (right).

Finally, most patients were taken multiple blood samples throughout their stay at the hospital. In total, there were 1523 blood samples with complete measurements of these three features for the training and test set of 375 patients plus 228 blood samples for the validation set of 29 patients. We validated our model with these additional blood tests (Supplementary Figure 4). Overall, the accuracy is 90%, further showing that the model can be applied to any blood sample, even this is far from a patient’s clinical outcome. The three selected features on survived and dead patients can be seen in Supplementary Figure 5. On average, the model could predict the outcome of the 26 validation patients from Tongji Hospital about 16 days in advance using all their blood samples (Figure 4), and it could predict the outcome of 377 (351+26) patients from Tongji Hospital in all training, test and validation sets about 9 days in advance using all available blood samples (Supplementary Figure 6).

**Figure 4:**
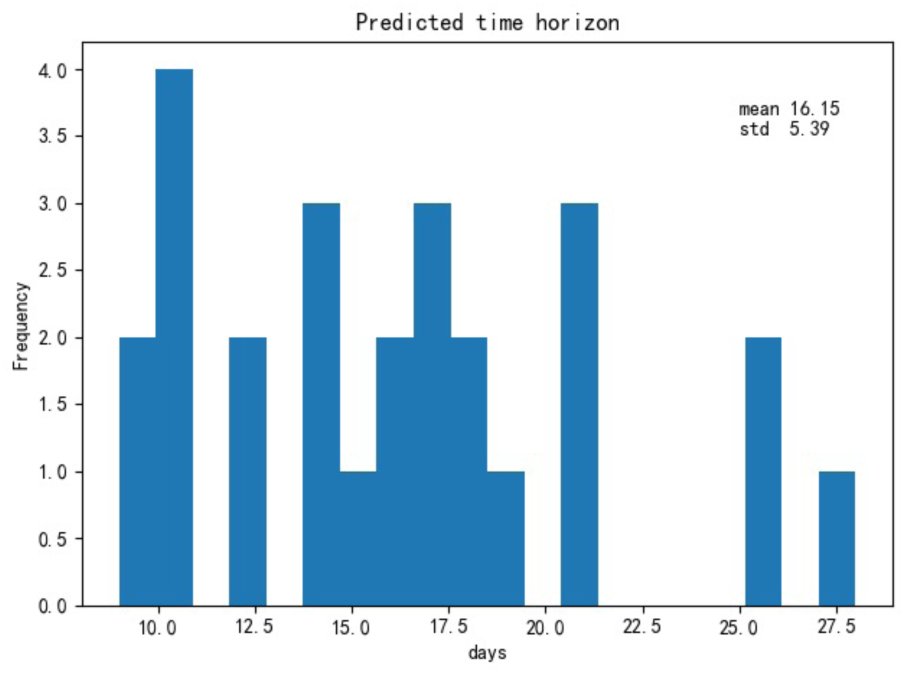
Histogram of the maximum number of days containing only true positive predictions until clinical outcome for 26 severe patients in the validation set (Tongji Hospital).

## Discussion

The significance of our work is two-fold. First, it goes beyond providing high-risk factors^2^. It provides a simple and intuitive clinical test to precisely and quickly quantify the risk of death. For example, a routine sequential respiratory support therapy for patients with SPO2 below 93% is: intranasal catheterization of oxygen, oxygen supply through mask, high flow oxygen supply through nasal catheter, non-invasive ventilation support, invasive ventilation support, and ECMO. Predicting that for some patients this sequential oxygen therapy leads to unsatisfactory therapeutic effects could pre-empt physicians to pursuit different approaches.

The goal is for the model to identify high-risk patients before irreversible lesions occur. Second, the three key features, LDH, lymphocytes and hs-CRP, can easily be collected by any hospital. In crowed hospitals, and with shortage of medical resources, this simple model can help to quickly prioritise patients.

The increase of LDH reflects tissue/cell destruction and is regarded as a common sign of tissue/cell damage. Serum LDH has been identified as an important biomarker for the activity and severity of Idiopathic Pulmonary Fibrosis (IPF)^6^. In patients with severe pulmonary interstitial disease, the increase of LDH is significant and is one of the most important prognostic markers of lung injury^6^. For the critically ill patients with COVID-19, the rise of LDH level indicates an increase of the activity and extent of lung injury.

Higher serum hs-CRP could also be used to predict the risk of death in severe COVID-19 patients. The increase of hs-CRP, an important marker for poor prognosis in ARDS^7,8^, reflects the persistent state of inflammation^9^. The result of this persistent inflammatory response is large grey-white lesions in the lungs of patients with COVID-19 (what was seen in the autopsy)^10^. In the tissue section, a large amount of sticky secretion was also seen overflowing from the alveoli^10^.

Finally, our results also suggested that lymphocyte may serve as a potential therapeutic target. The hypothesis is supported by results of clinical studies^2,11^. Moreover, Lymphopenia is a common feature in patients with COVID-19 and might be a critical factor associated with disease severity and mortality^12^. Injured alveolar epithelial cells could induce the infiltration of lymphocytes, leading to persistent lymphopenia as SARS-CoV and MERS-CoV did, given that they share similar alveolar penetrating and antigen presenting cells (APC) impairing pathways^13,14^. A biopsy study has provided strong evidence that the counts of peripheral CD4 and CD8 T cells were substantially reduced, while their status was hyperactivated^15^. Also, Jing and colleagues reported the lymphopenia is mainly related to the decrease of CD4 and CD8 T cells^16^. Thus, it is likely that lymphocytes play distinct roles in COVID-19, which deserves further investigation.

## Conclusion

This study has room for further improvement. First, since the proposed machine learning method is purely data driven, our model may vary if starting from different datasets. As more data become available, the whole procedure can easily be repeated to obtain more accurate models. This is a single-centred, retrospective study, which provides a preliminary assessment of the clinical course and outcome of severe patients. Although the original database covered more than 3,000 patients, most clinical outcomes had not yet been released at the time of this study. We look forward to subsequent large sample and multi-centred studies. Second, although we had a pool of more than 80 clinical measurements, here our modelling principle is a trade-off between the minimal number of features and the capacity of good prediction, therefore avoiding overfitting. Finally, this study strikes a balance between model interpretability and improved accuracy. While clinical settings tend to prefer interpretable models, it is possible that a black box model may lead to improved performance.

The recent data base of retrospective blood samples collected from 404 patients infected by COVID-19 in the Tongji Hospital in Wuhan, China, was used to identify predictive and potentially life-saving discriminative biomarkers of patients within a critical condition range. Our state-of-the-art machine learning framework suggests that the disease severity can be accurately predicted using three biomarkers, therefore greatly reducing the space of clinical parameters to be monitored and the associated medical burden.

In summary, this study identified three indicators (LDH, hs-CRP, and lymphocytes), with thresholds (LDH: 365U/l, hs-CRP: 41.2mg/L, and lymphocytes%: 14.7%) for COVID-19 prognostic prediction. We developed an XGBoost machine learning-based prognostic model that can predict the survival rates of severe patients with more than 90% accuracy using the last sample and 90% from any other blood sample, enabling detection, early intervention and potentially reduction of mortality in high-risk patients with COVID-19. From a technical point of view, this work helps pave the way for using machine learning method in COVID-19 prediction and diagnosis in the triage of the large scale explosive epidemic COVID-19 cases.

## Data Availability

Data is available once the paper gets accepted. The Python code is available upon request from YY.

## Supplementary Information

In the Supplementary Information, we shall illustrate data analysis using a step-by-step procedure below:

**Step1**. <u>Obtain the Top10 features using 375 samples with all features:</u>

**Supplementary Figure 1:**
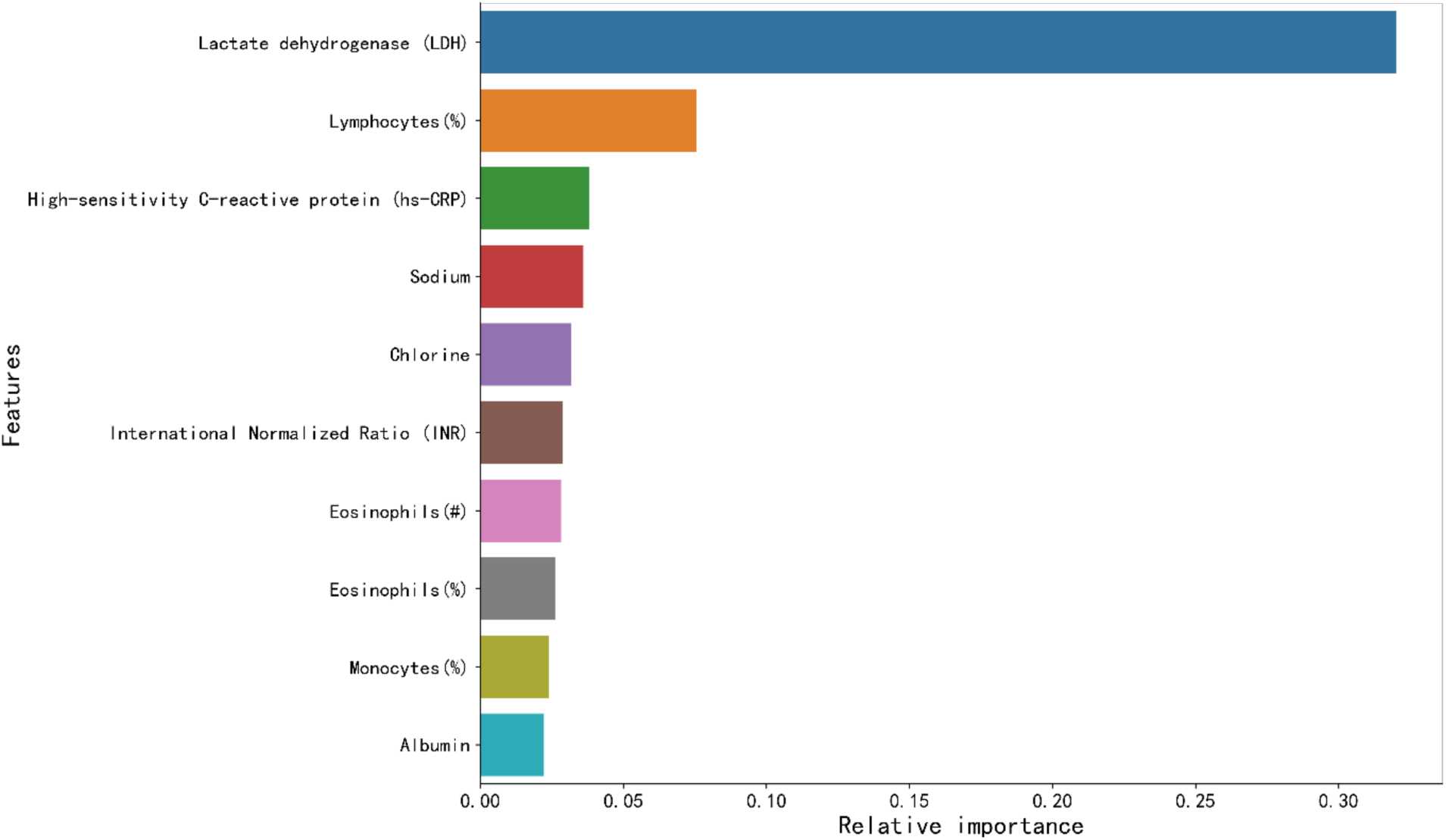
Top ten key clinical features that are ranked according to its importance in the Multi-tree XGBoost algorithm.

### Multi-tree XGBoost with 375 samples (all features)

It is trained with the parameters setting as the max depth with 4, the learning rate is equal 0.2, the tress number of estimators is set to 150, the value of the regularization parameter α is set to 1, the ‘subsample’ and ‘colsample_bytree’ both are set to 0.9 to prevent overfitting when there are many features and the sample size is not large.

**Step 2**. <u>Reduce the number of features used:</u>

Supplementary Algorithm 1:

#### Algorithm 1 Feature selection

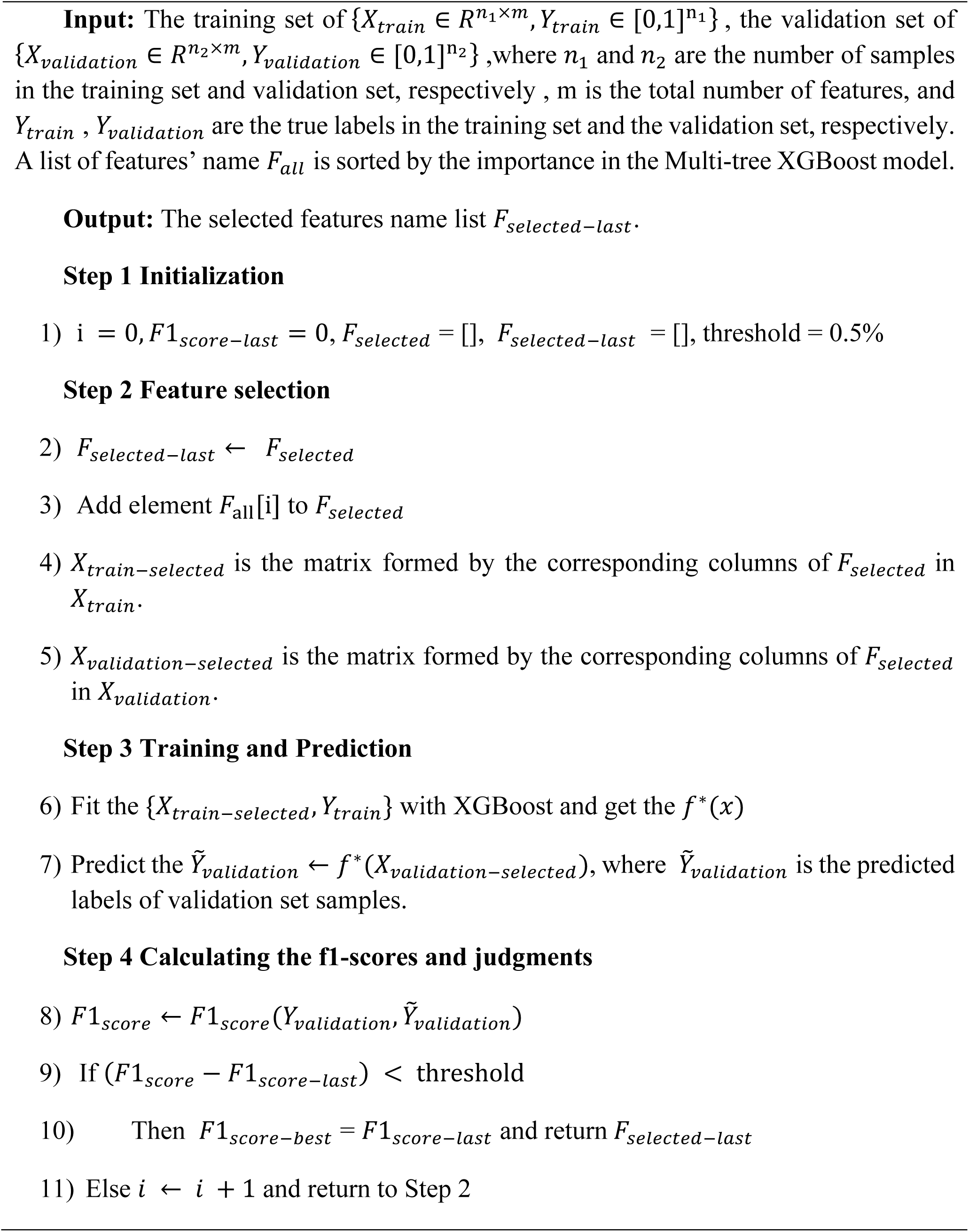

### Multi-tree XGBoost with 375 samples (Top-X features)

It is trained with the parameters setting as the max depth with 4, the learning rate is equal 0.2, the value of the regularization parameter α is set to 1.

**Supplementary Figure 2:**
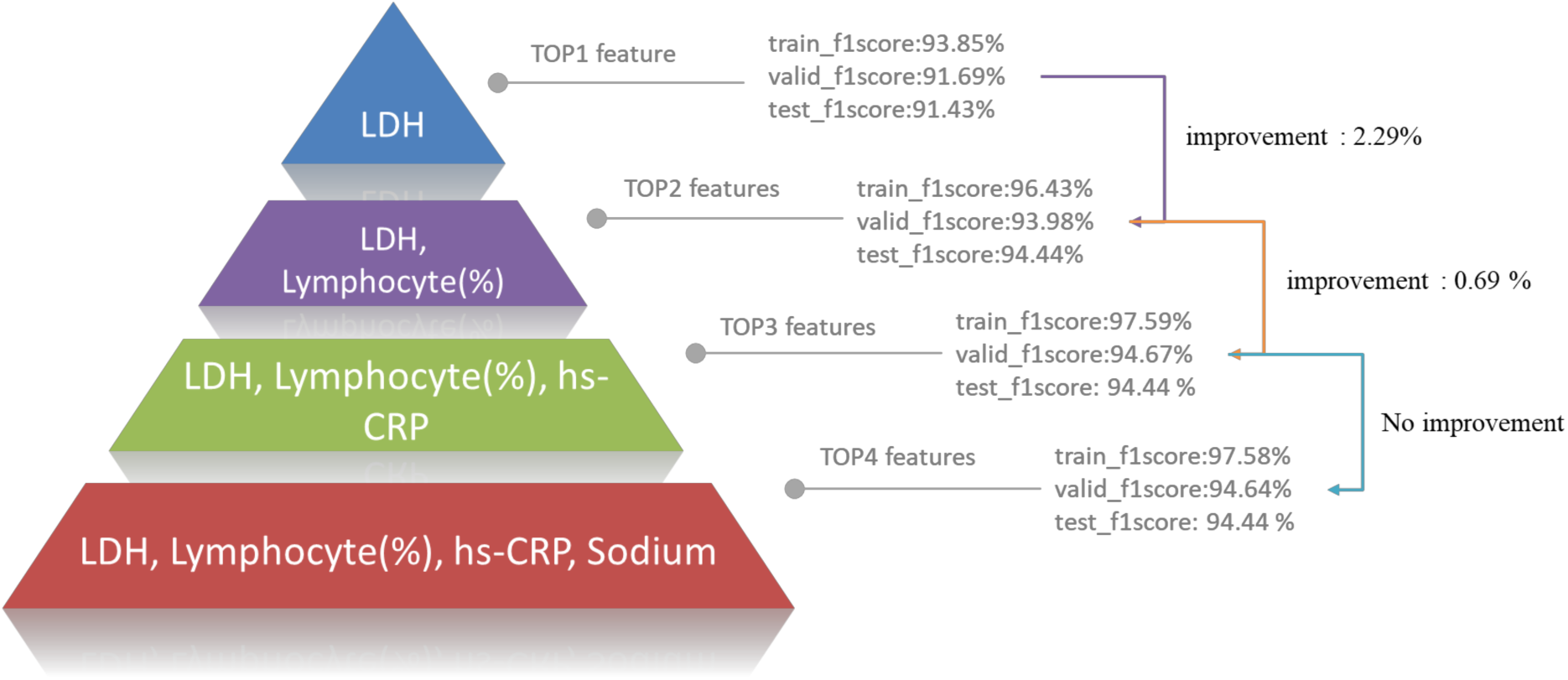
Illustration of F1 scores using Supplementary Algorithm 1.

**Step 3**. <u>The results on the Multi-tree XGBoost with Top 3 features selected in Step 2 (375 samples).</u>

**Supplementary Figure 3:**
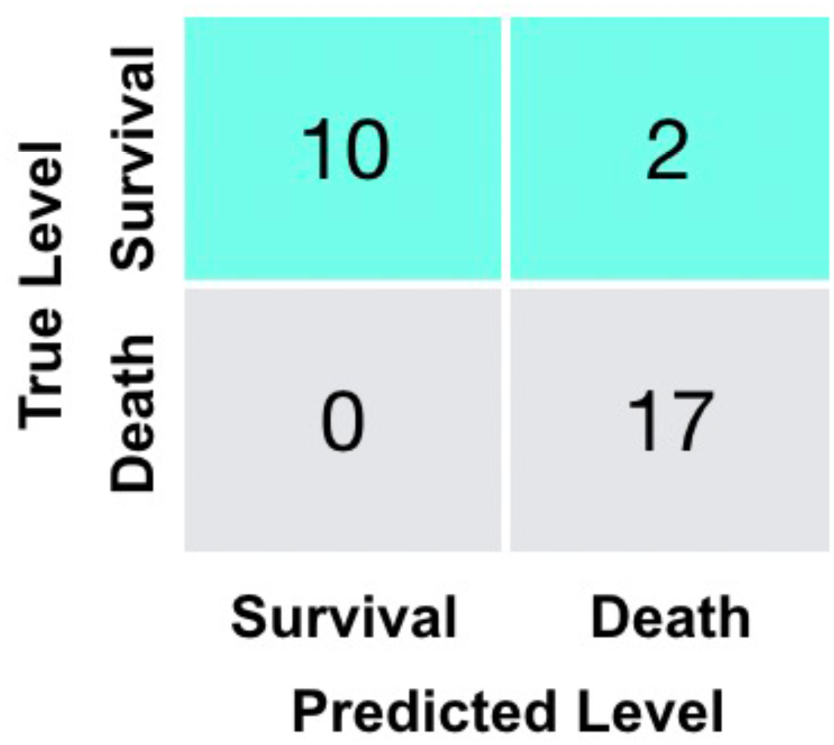
Confusion matrix for the additional validation dataset using the Multi-tree XGBoost algorithm.

**Supplementary Table 1.** Performance of the Multi-tree XGBoost algorithm on training dataset.

|  | **Precision** | **Recall** | **F1-score** | **Support** |
| --- | --- | --- | --- | --- |
| **Survival** | 1.00 | 0.95 | 0.98 | 145 |
| **Death** | 0.94 | 1.00 | 0.97 | 117 |
| **accuracy** |  |  | 0.97 | 262 |
| **macro avg** | 0.97 | 0.98 | 0.97 | 262 |
| **weighted avg** | 0.97 | 0.97 | 0.97 | 262 |

**Supplementary Table 2:** Performance of the Multi-tree XGBoost algorithm on testing dataset.

|  | **Precision** | **Recall** | **F1-score** | **Support** |
| --- | --- | --- | --- | --- |
| **Survival** | 0.96 | 0.95 | 0.95 | 56 |
| **Death** | 0.95 | 0.96 | 0.96 | 57 |
| **accuracy** |  |  | 0.96 | 113 |
| **macro avg** | 0.96 | 0.96 | 0.96 | 113 |
| **weighted avg** | 0.96 | 0.96 | 0.96 | 113 |

**Step 4**. <u>Reduce number of tree to 1, which leads to the Single-tree XGBoost algorithm. Because there are 24 samples with at least one of top-3 features missing. In order to obtain a better decision rule, we have deleted these samples and obtain a new dataset with the 351 samples and 3 features.</u>

### Single-tree XGBoost with 351 samples (Top3 features)

It is trained with the parameter setting as the tress number of estimators is set to 1, the values of the two regularization parameters α and β are both set to 0, the subsample and max features both are set to 1.

**Supplementary Table 3.** Performance of the proposed algorithm on training dataset for the Single-tree XGBoost algorithm.

|  | **Precision** | **Recall** | **F1-score** | **Support** |
| --- | --- | --- | --- | --- |
| **Survival** | 1.00 | 0.98 | 0.99 | 135 |
| **Death** | 0.97 | 1.00 | 0.99 | 110 |
| **accuracy** |  |  | 0.99 | 245 |
| **macro avg** | 0.99 | 0.99 | 0.99 | 245 |
| **weighted avg** | 0.99 | 0.99 | 0.99 | 245 |

**Supplementary Table 4:** Performance of the proposed algorithm on validation dataset for the Single-tree XGBoost algorithm.

|  | **Precision** | **Recall** | **F1-score** | **Support** |
| --- | --- | --- | --- | --- |
| **Survival** | 0.95 | 0.96 | 0.96 | 57 |
| **Death** | 0.96 | 0.94 | 0.95 | 49 |
| **accuracy** |  |  | 0.95 | 106 |
| **macro avg** | 0.95 | 0.95 | 0.95 | 106 |
| **weighted avg** | 0.95 | 0.95 | 0.95 | 106 |

**Supplementary Figure 4:**
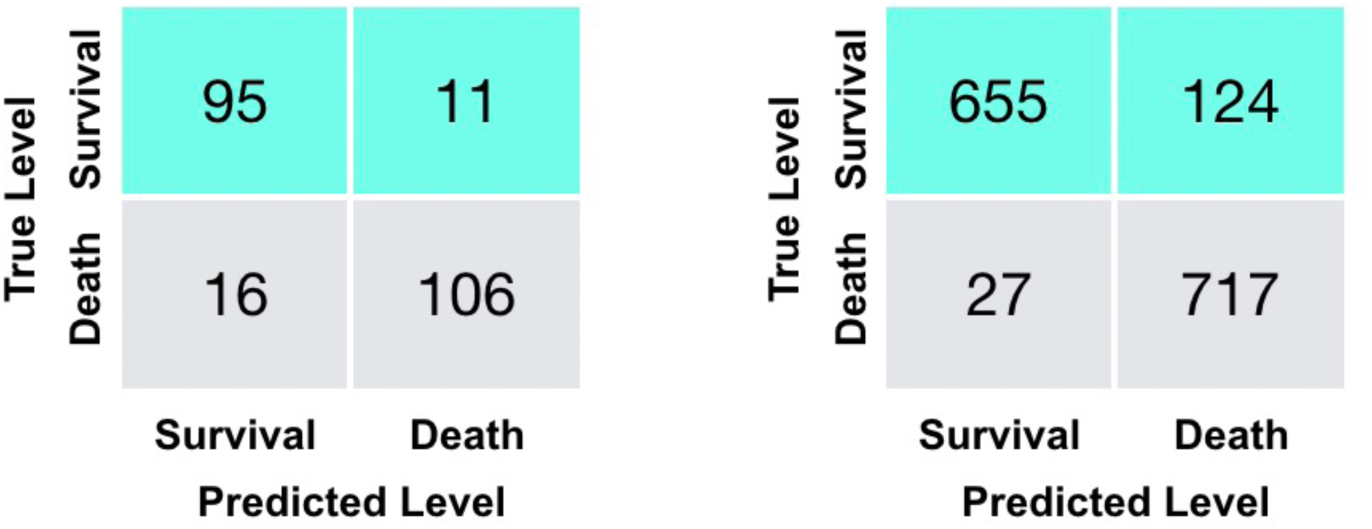
Confusion matrices for additional validation set (29 severe patients) with all 228 available blood samples (left), and training and testing sets (375 patients) with all 1523 available complete blood samples (right).

**Supplementary Figure 5:**
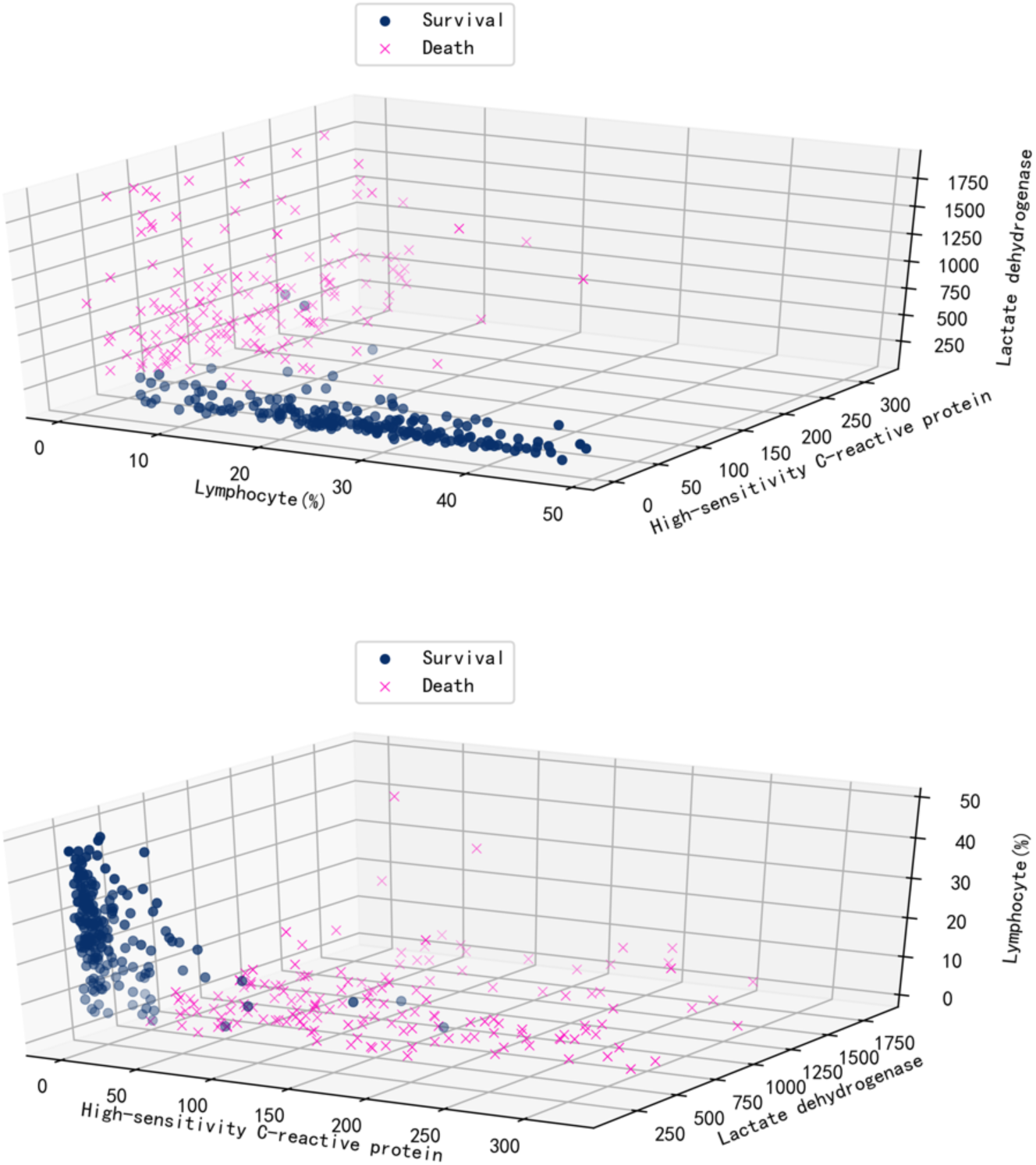
Visualization of data over three selected features.

**Supplementary Figure 6:**
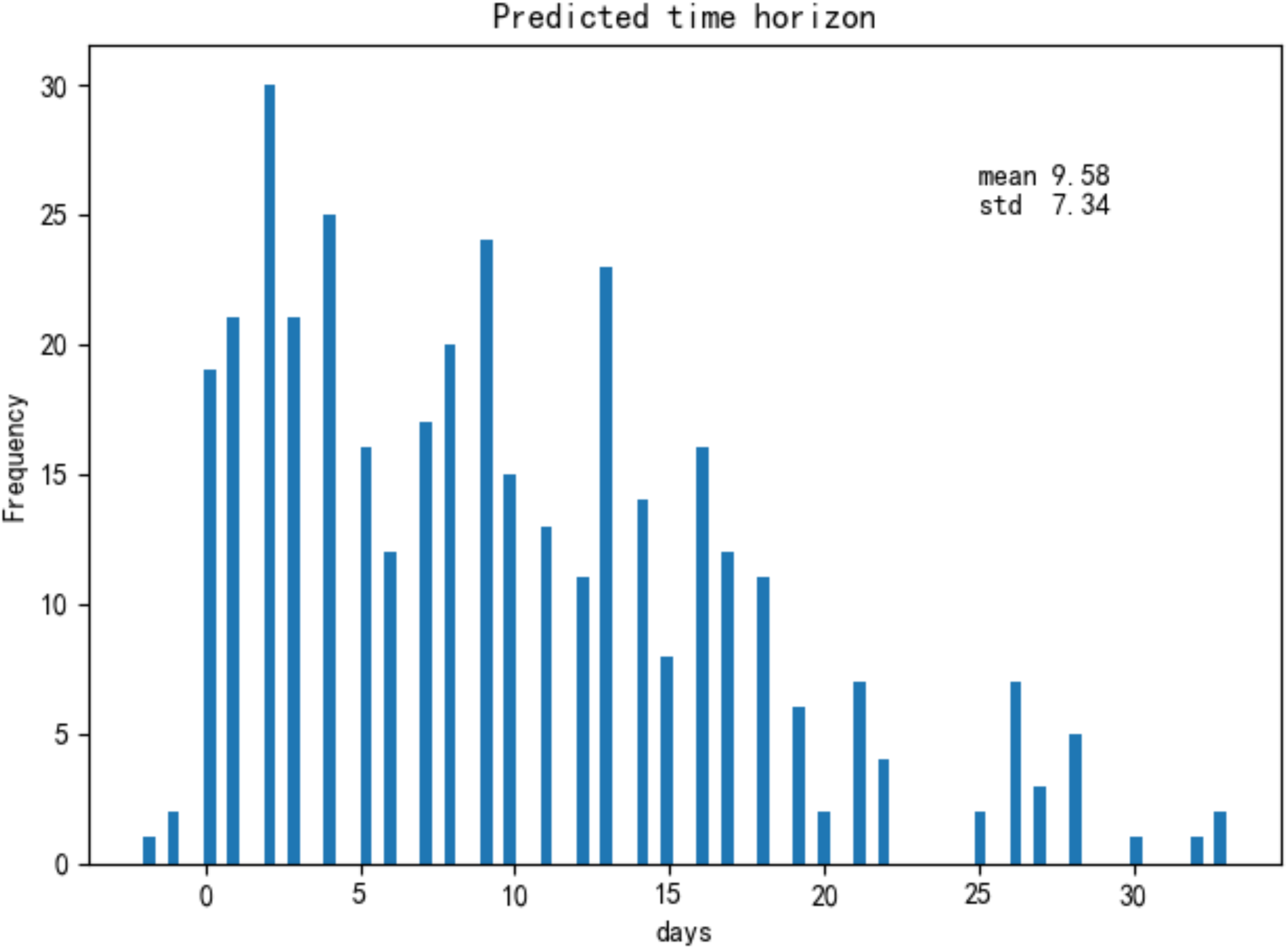
For 377 patients in both training and test sets, we plot the histogram of the number of days between their prediction and outcome. Note that there are three patients with negative days as their blood sample results arrived one and two days after their clinical outcome.

